# LDAK-GBAT: fast and powerful gene-based association testing using summary statistics

**DOI:** 10.1101/2022.07.01.22277161

**Authors:** Takiy-Eddine Berrandou, David Balding, Doug Speed

**Affiliations:** Quantitative Genetics and Genomics (QGG), Aarhus University, Aarhus, Denmark; Melbourne Integrative Genomics (MIG), Melbourne University, Melbourne, Australia

## Abstract

We present LDAK-GBAT, a novel tool for gene-based association testing using summary statistics from genome-wide association studies. We first evaluate LDAK-GBAT using ten phenotypes from the UK Biobank. We show that LDAK-GBAT is computationally efficient, taking approximately 30 minutes to analyze imputed data (2.9M common, genic SNPs), and requiring less than 10Gb memory. In total, LDAK-GBAT finds 680 genome-wide significant genes (*P*≤2.8×10^−6^), which is at least 25% more than each of five existing tools (MAGMA, GCTA-fastBAT, sumFREGAT-SKAT-O, sumFREGAT-PCA and sumFREGAT-ACAT), and 48% more than found by single-SNP analysis. We then analyze 99 additional phenotypes from the UK Biobank, the Million Veterans Project and the Psychiatric Genetics Consortium. In total, LDAK-GBAT finds 7957 significant genes, which is at least 24% more than the best existing tools, and 42% more than found by single-SNP analysis.

## INTRODUCTION

A genome-wide association study (GWAS) analysis typically starts by testing each SNP individually for association with the phenotype ^1^. However, it is now common to also perform a gene-based analysis, which instead tests for association sets of SNPs defined by gene annotations ^2,3^. Gene-based analyses can be more powerful than single-SNP analyses because they are able to accumulate evidence of association across multiple SNPs, and because they perform fewer statistical tests (and so require less correction for multiple testing) ^4-6^. Further, gene-based analyses are biologically justified, as it is plausible that variants within a gene act jointly to affect an outcome ^4^.

We propose LDAK-GBAT, a tool for performing gene-based association analysis. LDAK-GBAT extends a previous tool, FaST-LMM-Set ^7^, in three key ways. Firstly, LDAK-GBAT can be applied using GWAS summary statistics and a reference panel, whereas FaST-LMM-Set requires individual-level data. Secondly, LDAK-GBAT accommodates alternative heritability models (assumptions regarding how heritability is distributed across the genome), whereas FaST-LMM-Set assumes that all SNPs are expected to contribute equally towards the phenotype. Thirdly, results from LDAK-GBAT can be clumped, which provides an approximate way to divide the significant genes into those directly and indirectly associated with the phenotype.

First, we analyze summary statistics for ten phenotypes from UK Biobank (UKBB) ^8^. We show that LDAK-GBAT is computationally efficient, produces well-calibrated p-values under the null hypothesis, and finds at least a quarter more significant genes than MAGMA ^9^, GCTA-fastBAT ^10^, sumFREGAT-SKAT-O, sumFREGAT-PCA, sumFREGAT-ACAT ^11,12^ and single-SNP analysis. Next, we analyze summary statistics for 99 traits: 72 additional phenotypes from UK Biobank defined by the International Classification of Diseases 10th Revision (ICD-10), 18 traits from Million Veterans Project (MVP) ^13^ and nine traits from the Psychiatric Genomics Consortium (PGC) ^14^. Again, LDAK-GBAT finds substantially more significant genes than MAGMA, sumFREGAT-PCA, sumFREGAT-ACAT and single-SNP analysis.

LDAK-GBAT is freely-available within our software package LDAK (see Web Resources for a link to the LDAK website, that contains full documentation, including test data and example scripts).

## MATERIAL AND METHODS

Here we summarize how LDAK-GBAT tests genes for association with a phenotype; for full details see the Supplementary Note. Suppose we have a GWAS of n individuals, and we wish to test a gene containing m SNPs. Let the length-n vector *Y* contain the phenotypic values, and the *n* × *m* matrix *X* contain the SNP genotypes. For convenience, we assume that *X* _*j*_, the jth column of *X*, has been standardized to have mean zero and variance one. LDAK-GBAT assumes the linear model

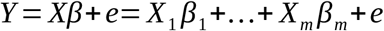

Here, *β* _*j*_ denotes the effect size for the jth SNP, while the vector e denotes environmental noise. We assign the following prior distributions to *β* _*j*_ and e

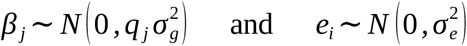

where 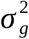 and 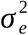 denote genetic and environmental variance components, respectively, and the *q* _*j*_ are pre-specified constants determined by the choice of heritability model (see below). We estimate 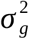 and 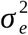 using REML (restricted maximum likelihood) ^15^, then obtain p-values via a likelihood ratio test of 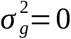. Like FaST-LMM-Set, we derive the null distribution of the test statistic via permutation ^7^.

As well as p-values, LDAK-GBAT reports *X* 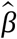 for each gene, an estimate of its genetic contribution (in effect, a polygenic risk score computed using only the SNPs in the gene). These estimates can be used to clump the results from LDAK-GBAT (similar to how it is standard to clump the results from single-SNP analysis ^16^). For example, in the analyses below, we compute the squared correlation between the estimated genetic contributions of significant genes on the same chromosome; if a pair of genes has squared correlation above 0.1, we exclude the gene with highest p-value.

The above description assumes we have individual-level data (access to *X* and *Y*). However, in the Supplementary Note, we explain how all the required calculations can be written in terms of the form *Cor* (*X* _*j*_, *Y*), the correlation between SNP j and the phenotype, and *Cor* (*X* _*j*_, *X*_*k*_), the correlation between SNPs j and k. This enables LDAK-GBAT to be run using only summary statistics from single-SNP analysis (from which it recovers *Cor* (*X* _*j*_, *Y*) for each SNP) and a reference panel (which it uses to estimate *Cor* (*X* _*j*_, *X*_*k*_) between pairs of SNPs).

When analyzing SNP data, the heritability model describes how 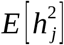, the expected heritability contributed by a SNP, varies across the genome. With SNPs standardized, 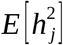 is proportional to 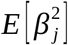, which is in turn proportional to *q* _*j*_ (i.e., the values for *q* _*j*_ determine the heritability model). If we set *q* _*j*_=1 for all SNPs, then LDAK-GBAT assumes the same model as FaST-LMM-Set. This choice of *q* _*j*_ corresponds to assuming 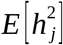 is constant, and therefore we refer to it as the “Uniform Model”. Our previous works indicate that the Uniform Model is sub-optimal, because when analyzing real data, we observed that 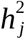 systematically varies with features such as minor allele frequency (MAF) and linkage disequilibrium (LD)^17-20^.

### UKBB, MVP and PGC summary statistics

We compute summary statistics for the UKBB phenotypes from the individual-level data. We first focus on ten phenotypes: body mass index, college education, forced vital capacity, height, hypertension, impedance, neuroticism score, preference for evenings, pulse rate, and systolic blood pressure. We subsequently consider 72 ICD-10 diseases, spanning 12 of the 22 chapters (see Supplementary Table 1 for details). We compute summary statistics by performing classical linear regression (for both quantitative and binary traits), restricting to unrelated, white British individuals, and including 13 covariates (age, sex, Townsend Deprivation Index and ten principal components). For the first ten phenotypes, the sample size is 50,000 individuals (the exception is when performing single-SNP analysis, where we use up to 200,000 individuals). For the ICD-10 phenotypes, there are on average 12,781 cases (range 1,559 to 70,012) and 225,130 controls (range 103,972 to 250,346). Further details of the UKBB summary statistics are provided in the Supplementary Note.

For MVP and PGC, we use summary statistics from previously published GWAS. For MVP, we consider 18 phenotypes including type 2 diabetes, cardiovascular diseases, number of cigarettes per day, lipid levels and blood pressure The average sample size is 242,207 (range 17,014 to 1,114,458). For PGC, we consider 9 phenotypes: alcohol use, Alzheimer’s Disease, attention deficit hyperactivity disorder, autism spectrum disorder, bipolar disorder, eating disorder, major depressive disorder, post-traumatic stress disorder and schizophrenia. The average sample size is 256,250 (range 46,351 to 762,917). See Supplementary Tables 2 and 3 for more details.

### Default settings of LDAK-GBAT

When running LDAK-GBAT, the user must choose a reference panel, provide gene annotations and specify a heritability model. For our analyses below, our primary reference panel comprises 10,000 unrelated UKBB individuals genotyped for 7,186,768 imputed SNPs with MAF ≥ 0.01 and imputation quality information (Rsq) ≥ 0.8. We use RefSeq gene annotations ^21^. When testing a gene, we consider only SNPs located between the transcription start and stop sites. In total, there are 17,965 genes, that contain 2,890,640 of the 7,186,768 SNPs. Unless stated otherwise, we run LDAK-GBAT assuming the “Human Default Model”. This sets *q*_*j*_ = [*p*_*j*_ (1− *p*_*j*_)]^0.75^, where *p* _*j*_ is the MAF of SNP j, and is our default recommendation when analyzing human traits ^19,20^.

### Existing tools

We compare LDAK-GBAT with five existing summary statistic tools for gene-based association analysis: MAGMA, GCTA-fastBAT. sumFREGAT-SKAT-O, sumFREGAT-PCA and sumFREGAT-ACAT (see Supplementary Note for a brief description of each tool). We use the default settings for each tool. MAGMA, GCTA-fastBAT, sumFREGAT-SKAT-O and sumFREGAT-PCA require a reference panel, for which we use the same 10,000 unrelated UKBB individuals that we use when running LDAK-GBAT.

## RESULTS

Supplementary Figures 1 and 2 show that when we run LDAK-GBAT assuming the Uniform Model, its results align closely with those from FaST-LMM-Set, confirming that it is feasible to use only GWAS summary statistics and a reference panel, instead of individual-level data.

### Comparing LDAK-GBAT with existing tools

Firstly, we compare tools based on their type 1 error. For this we generate permuted versions of the UKBB phenotype height. If a tool effectively controls the type 1 error, then the p-values from analyzing the permuted phenotype should be uniformly distributed. Supplementary Table 4 shows that this is the case for LDAK-GBAT, and is generally the case for the five existing tools. For example, for LDAK-GBAT, we find that 0.0099, 1.0×10^−4^ and 1.5×10^−6^ genes have p-values less than 0.01, 1×10^−4^ and 1×10^−6^, respectively.

Secondly, we compare tools based on computational efficiency. Table 1 shows that sumFREGAT-ACAT, LDAK-GBAT and MAGMA are the fastest tools, taking on average 20-50 minutes to analyze each of the ten UKBB phenotypes. Meanwhile, sumFREGAT-PCA and sumFREGAT-SKAT-O are the slowest tools, taking over 10 hours to analyze each phenotype. All tools have either low or modest memory demands, requiring at most 23Gb.

**Table 1:** Computational efficiency of different tools for gene-based association testing. Average time and memory required to analyze each of the ten UKBB phenotypes on a single CPU.

| Tool | Time (minutes) | Memory (GB) |
| --- | --- | --- |
| LDAK-GBAT | 33 | 5 |
| MAGMA | 47 | 1 |
| GCTA-fastBAT | 96 | 22 |
| sumFREGAT-SKAT-O | 1299 | 23 |
| sumFREGAT-PCA | 679 | 18 |
| sumFREGAT-ACAT | 20 | 9 |

Thirdly, we compare tools based on the number of significant genes. We use the Bonferroni significance threshold 0.05/17669 = 2.8×10^−6^. Figure 1 and Supplementary Table 5 show that, in total, LDAK-GBAT finds 680 significant genes, whereas MAGMA, GCTA-fastBAT, sumFREGAT-SKAT-O, sumFREGAT-PCA and sumFREGAT-ACAT find 410, 408, 93, 542 and 534, respectively. Supplementary Tables 5 and 6 show that LDAK-GBAT continues to find the most associations if we filter significant genes based on LD, or if we restrict our analysis to 606k directly genotyped SNPs.

**Figure 1:**
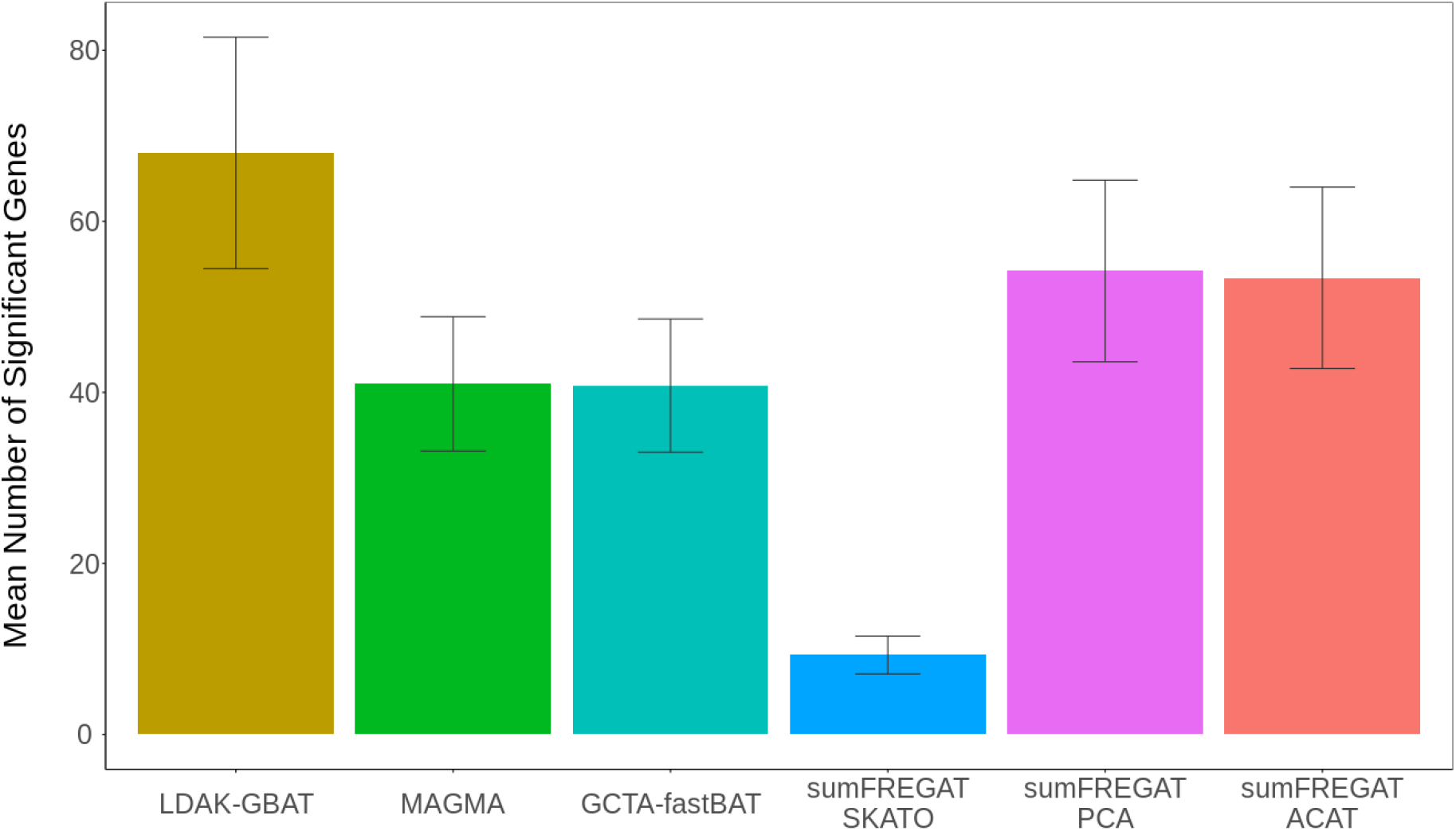
Comparing gene-based association testing tools for the ten UK Biobank phenotypes. Bars report the mean number of genome-wide significant genes (P≤2.8×10^−6^) for LDAK-GBAT and five existing tools. Segments mark 95% confidence intervals for the means.

### Comparing LDAK-GBAT with single-SNP analysis

When performing single-SNP analysis, we declare a gene significant if it contains at least one SNP with *P*≤5e-8. Figure 2 shows that when analyzing 50,000 individuals, single-SNP analysis finds in total 460 significant genes, 220 fewer than LDAK-GBAT. Further, we see that for single-SNP analysis to find the same number of significant genes as LDAK-GBAT, we must increase the sample size to approximately 65,000 individuals (i.e., by about 30%). Supplementary Table 7 shows that 249 of the 267 genes (93%) significant from LDAK-GBAT but not from single-SNP analysis, are significant from single-SNP analysis when the sample size is increased to 200,000 individuals, providing reassurance that the majority of novel associations found by LDAK-GBAT are true positives.

**Figure 2:**
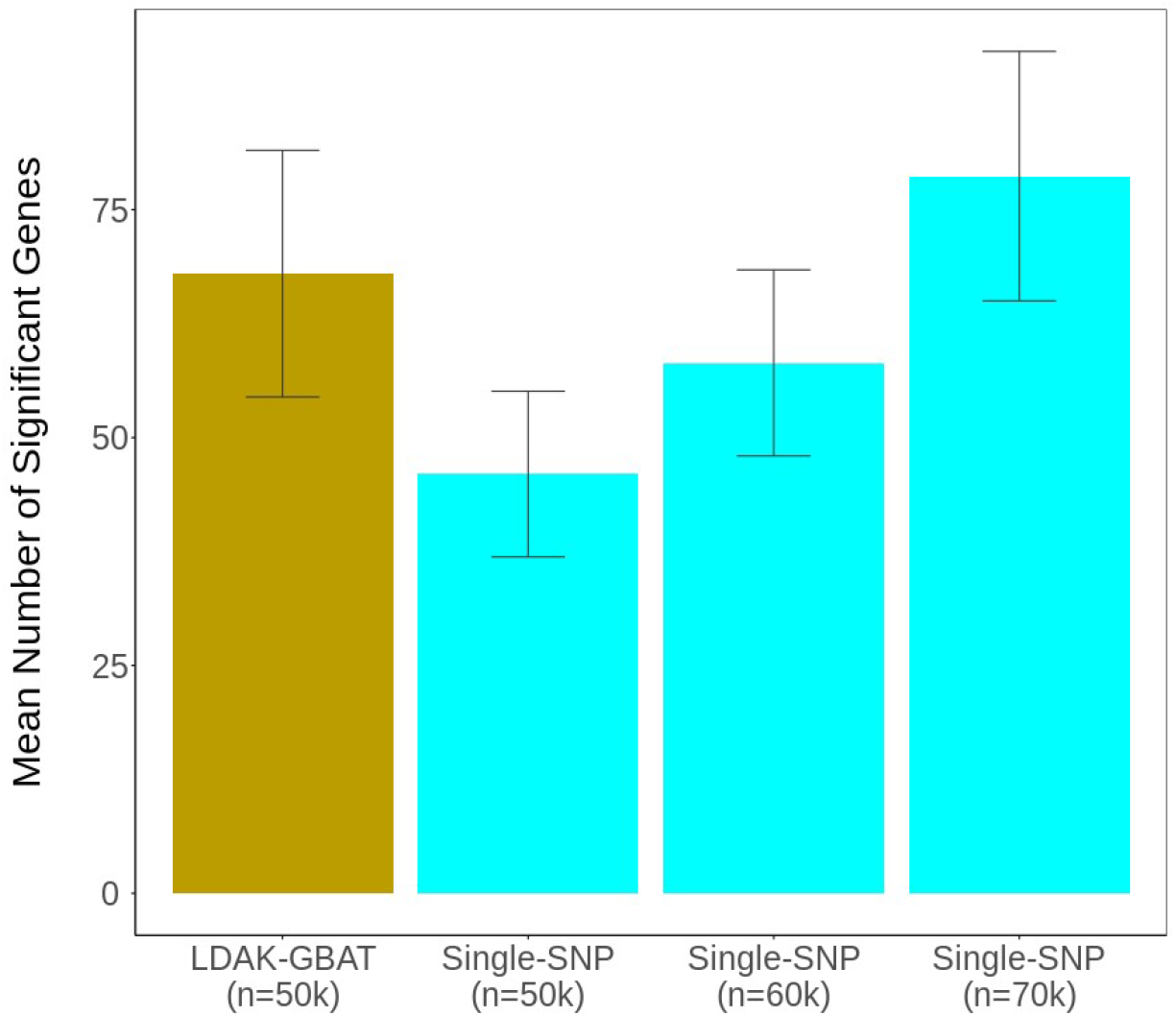
Comparing LDAK-GBAT and single-SNP analysis for the ten UK Biobank phenotypes. Bars report the mean number of genome-wide significant genes (P≤2.8×10^−6^) for LDAK-GBAT (using 50,000 individuals) and the number of genes containing a SNP with P≤5e-8 from single-SNP analysis (using 50,000, 60,000 or 70,000 individuals). Segments mark 95% confidence intervals for the means.

### Choice of reference panel and heritability model

Supplementary Figure 3 shows that results from LDAK-GBAT are very similar if we replace the UKBB reference panel with 404 non-Finnish, European individuals from the 1000 Genome Project ^21^. Figure 3 and Supplementary Table 8 consider heritability models of the form *q* _*j*_ = [*p* _*j*_ (1− *p*_*j*_)]^1+*α*^ for α = −1.25, −1, −0.75, −0.5, −0.25, 0 and 0.25. We see that changing from the Uniform Model (α=-1) to the Human Default Model (α=-0.25) has increased the total number of significant genes from 532 to 680 (27%).

**Figure 3:**
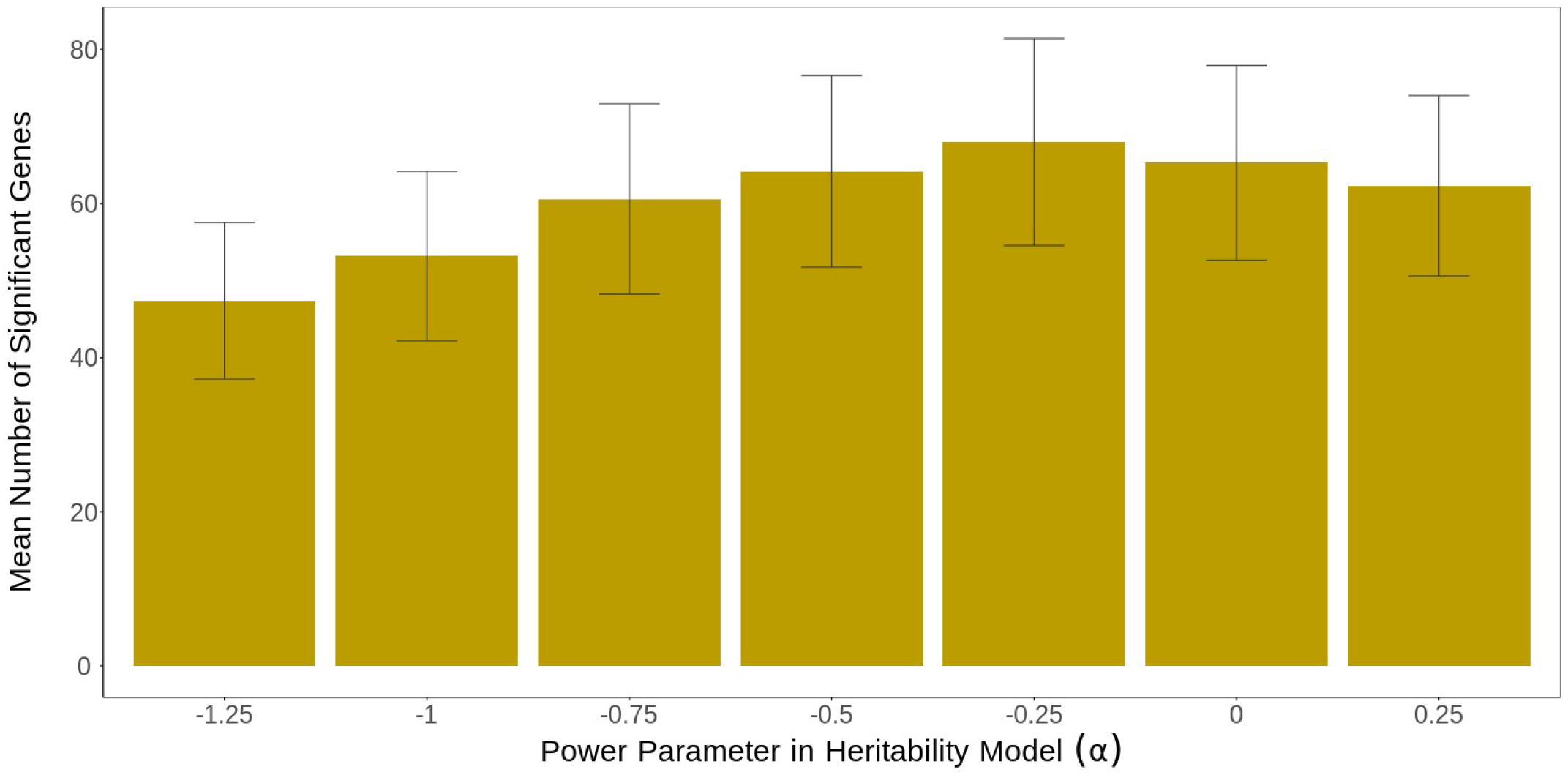
Varying the heritability model for the ten UK Biobank phenotypes. We run LDAK-GBAT using seven heritability models, defined by 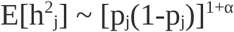, where p_j_ is the MAF of SNP_j_, and α is −1.25, −1, - 0.75, −0.5, −0.25, 0 or 0.25. Bars report the mean number of genome-wide significant genes (P≤2.8×10^−6^). Segments mark 95% confidence intervals for the means.

### Analysis of ICD-10, MVP and PGC phenotypes

Figure 4 and Supplementary Table 9 report results from analyzing the 99 ICD10, MVP and PGC phenotypes. Now we compare LDAK-GBAT with MAGMA, sumFREGAT-PCA and sumFREGAT-ACAT (the three best-performing existing tools when analyzing the ten UKBB phenotypes). We continue to use the Bonferroni significance threshold 2.8e-6. Across the 72 UKBB ICD10 phenotypes, LDAK-GBAT finds 1874 significant genes, whereas MAGMA, sumFREGAT-PCA and sumFREGAT-ACAT find only 1265, 1560 and 1609, respectively. The pattern is similar for the 18 MVP traits (LDAK-GBAT finds 4681 significant genes, compared to 2964, 3456 and 3659), and also for the nine PGC traits (LDAK-GBAT finds 1402 significant genes, compared to 943, 1171 and 1130). We note that 1279 (16%) of the significant genes from LDAK-GBAT are not detected by any of the three existing tools, nor by single-SNP analysis (Supplementary Figure 4).

**Figure 4:**
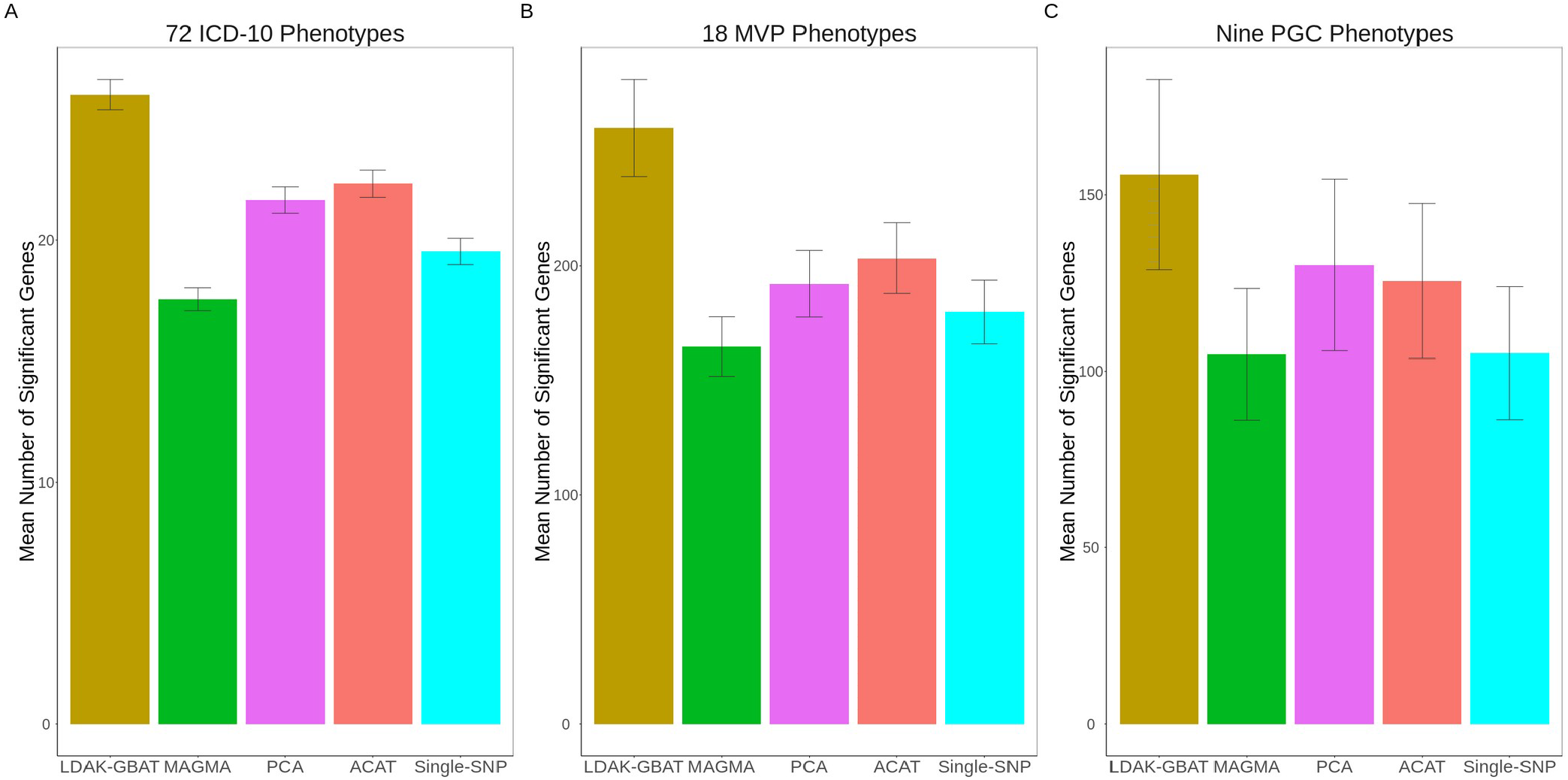
Comparing gene-based association testing tools for the 99 ICD10, MVP and PGC phenotypes. Bars report the mean number of genome-wide significant genes (P≤2.8×10^−6^) for LDAK-GBAT and three existing tools, and the number of genes with a SNP with P<5e-8 from single-SNP analysis, across the 72 ICD10 phenotypes (left), the 18 MVP phenotypes (middle) and nine PGC phenotypes (right). Segments mark 95% confidence intervals for the means.

For Supplementary Table 10, we clump the results from LDAK-GBAT so that no two genes on the same chromosome have estimated genetic contributions with squared correlation greater than 0.1. This reduces the 7957 significant genes to 2290 approximately independent significant genes (448, 1362 and 480 for the ICD10, MVP and PGC traits, respectively). To illustrate some of the benefits of clumping, Figure 5 compares results from LDAK-GBAT and single-SNP analysis for Type 2 Diabetes (the trait with most significant genes), for Chromosome 3 45 - 90 Mbp. In total, LDAK-GBAT finds that 50 of 261 genes in this region are significantly associated. However, clumping reduces this to ten approximately independent significant genes. For example, there are 39 significant genes in the first 10Mbp of the region, however, clumping indicates that 37 of these are associated only because they are in LD with the genes RBM6 or SFMBT1. Similarly, there are ten significant genes in the last 5Mbp of the region, however, clumping indicates that nine of these are associated only because they are in LD with the gene VGLL3. Moreover, it is interesting to note that VGLL3 is strongly significant from LDAK-GBAT (*P*=3.8×10^−8^), but only modestly significant from single-SNP analysis (across the 43 SNPs within this gene, the smallest *p*-value from single-SNP analysis is 7×10^−5^).

**Figure 5:**
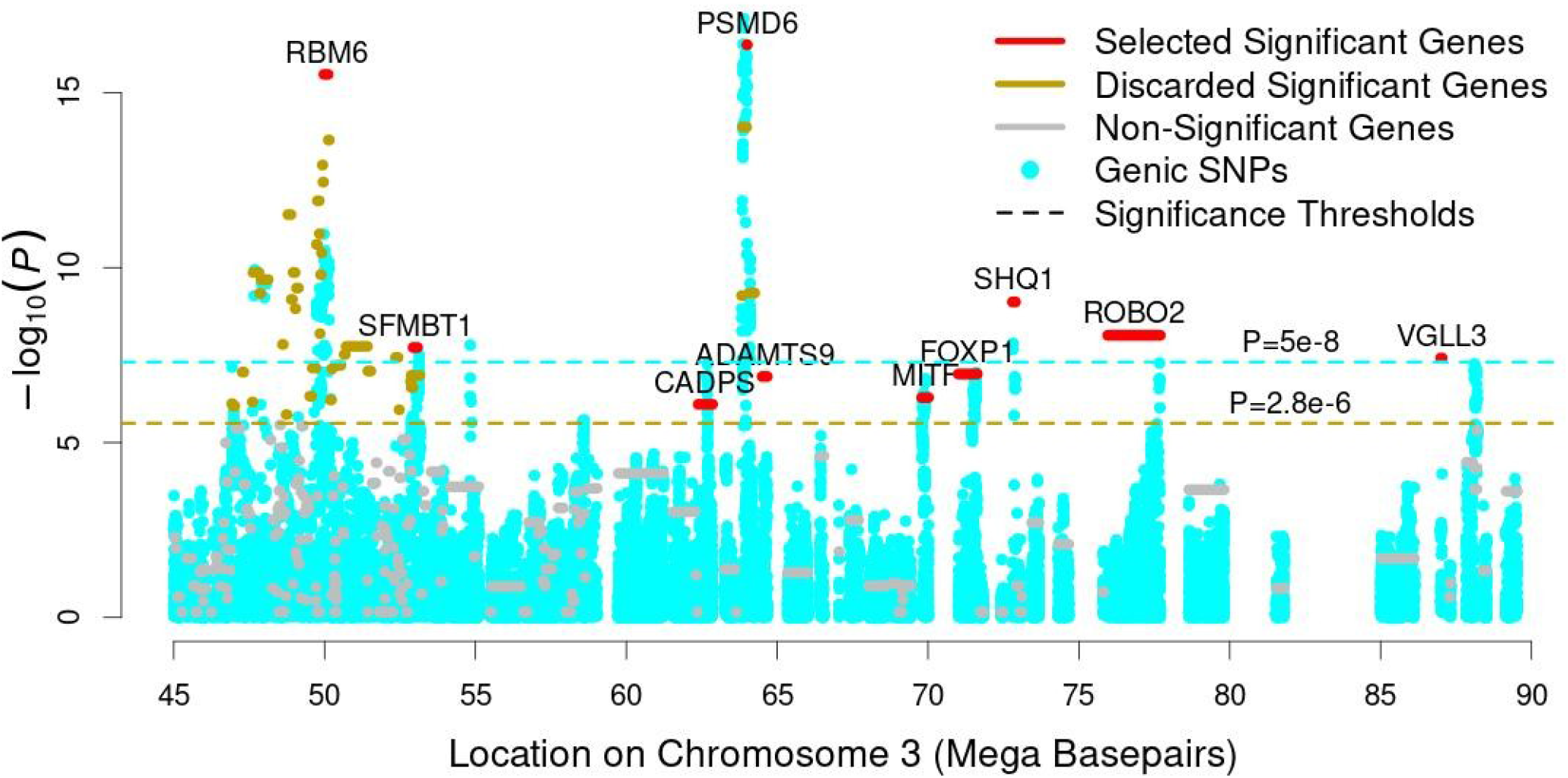
Clumping results from LDAK-GBAT. Segments provide p-values from LDAK-GBAT when analyzing the MVP phenotype type 2 diabetes; red segments indicate significant genes that are selected by clumping, gold segments indicate significant genes that are discarded by clumping, while gray segments indicate non-significant genes. Blue points report p-values for genic SNPs from single-SNP analysis. The gold and blue horizontal lines mark P=2.8×10^−6^ and P=5×10^−8^, respectively (the significance thresholds used with LDAK-GBAT and single-SNP analyses).

## DISCUSSION

We have proposed LDAK-GBAT, a novel method for gene-based association analysis. We have shown that LDAK-GBAT is both computationally efficient and statistically more powerful than existing tools for gene-based association testing.

LDAK-GBAT improves the existing tool FaST-LMM-Set in three main ways. Firstly, LDAK-GBAT uses GWAS summary statistics and a reference panel, whereas FaST-LMM-Set requires individual-level data. This made it feasible, for example, for us to analyze phenotypes from the MVP and PGC without access to individual-level data. Secondly, LDAK-GBAT allows the user to specify the heritability model, whereas FaST-LMM-Set considers only the Uniform Model. Figure 3 showed that, for the ten UKBB phenotypes, switching from the Uniform Model to the Default Human Model resulted in 27% more significant genes (approximately equal to power difference between LDAK-GBAT and sumFREGAT-PCA, the best existing tool). We speculate that it may be possible to increase the power of LDAK-GBAT further by considering more complex heritability models, that take into account, for example, functional annotations ^19,20^. Thirdly, results from LDAK-GBAT can be clumped. This feature is particularly useful when analyzing results from large GWAS (e.g., >100,000 samples), where it is common to find clusters of significantly associated genes, and so clumping provides a quick way to prioritize the genes in each cluster, and to estimate the number of independent significant associations.

We recognize three limitations with the LDAK-GBAT methodology. Firstly, the accuracy of LDAK-GBAT depends on the suitability of the reference panel. In this respect, it is reassuring that when analyzing the ten UKBB phenotypes, results from LDAK-GBAT were robust to replacing the UKBB reference panel with 404 European individuals from the 1000 Genome Project (i.e., both reducing the size of the reference panel, and reducing its ancestral similarity with the GWAS data). Secondly, the present version of LDAK-GBAT can only be applied to results from single-ancestry GWAS, which is problematic considering the increasing popularity of multi-ancestry GWAS ^22^. As a possible solution, we will explore whether it is feasible to meta-analyze results from LDAK-GBAT across ancestries or develop a version of LDAK-GBAT that uses a multi-ancestry reference panel. Thirdly, although we believe that LDAK-GBAT is the first gene-based association tool to offer a clumping feature, we appreciate that this is an imperfect way to prioritize results (it assumes that the causal gene has the smallest p-value). We note that when performing single-SNP analyses, there are now a variety of fine-mapping methods that are more effective than clumping, and we believe that LDAK-GBAT will enable similar methods to be developed for gene-based analyses.

## Data Availability

This study used only previously-collected data, that are either freely available or can be applied for. Specifically, UK Biobank data can be applied for from www.ukbiobank.ac.uk. website, Million Veterans Project summary statistics can be applied for from https://www.ncbi.nlm.nih.gov/gap, and Psychiatric Genomics Consortium summary statistics can be downloaded from https://www.med.unc.edu/pgc/download-results.

## Appendices

None

## Description of supplementary material

The supplementary material include three notes, six figures and ten tables.

## Acknowledgments

T-E.B. and D.S. are supported by Aarhus University Research Foundation (AUFF), by the Independent Research Fund Denmark (Project no. 7025-00094B), and by a Lundbeck Foundation Experiment Grant. D.B. and D.S. are supported from the Australian Research Council (Grant DP190103188).

## Author contributions

Writing and editing the manuscript: T-E.B., D.B. and D.S. Study design/conception: T-E.B., D.B. and D.S. Analyses: T-E.B. and D.S.

## Web resources

LDAK-GBAT, https://www.ldak.org

MAGMA, https://ctg.cncr.nl/software/magma

GCTA-fastBAT, https://yanglab.westlake.edu.cn/software/gcta/#fastBAT

sumFREGAT, https://cran.r-project.org/web/packages/sumFREGAT

Psychiatric Genomics Consortium, https://www.med.unc.edu/pgc/download-results

dbGaP, https://www.ncbi.nlm.nih.gov/gap

TB’s GitHub, https://github.com/takiy-berrandou/LDAK-GBAT-paper-scripts

## Data and code availability

Instructions for running LDAK-GBAT are available on the LDAK website, while sample code for the analyses in this paper are on the GitHub page of TB. We applied for and downloaded individual-level UK Biobank data (via Application 21432) from the UK Biobank website. We applied for study acces-sion phs001672.v7.p1 and downloaded summary statistics for the Million Veterans Project GWAS from the dbGaP website. We downloaded summary statistics for the Psychiatric Genomics Consortium from the consortium’s website. See Web resources for links to each website.

