## Supplementary Notes and Figures for "LDAK-GBAT: fast and powerful gene-based association testing using summary statistics"

This file contains three supplementary notes and six supplementary figures. Note that ten supplementary tables are provided in a separate excel file.

**Supplementary notes**

Supplementary Note 1: Mathematical details of LDAK-GBAT
Supplementary Note 2: Existing summary statistic tools for gene-based association testing
Supplementary Note 3: Further details of data

**Supplementary figures**

Supplementary Figure 1:  Comparison of FaST-LMM-Set and LDAK-GBAT.
Supplementary Figure 2:  Comparison of LDAK-GBAT using individual-level data and summary statistics.
Supplementary Figure 3: Impact of changing the reference panel.
Supplementary Figure 4: Overlap between significant genes from different tools.
Supplementary Figure 5: The impact of pruning genes.
Supplementary Figure 6: Impact of number of permutations.

**Supplementary tables (excel file)**

Supplementary Table 1: Details of the ICD10 phenotypes.
Supplementary Table 2: Details of the Millions Veterans Project phenotypes.
Supplementary Table 3: Details of the Psychiatric Genomics Consortium phenotypes.
Supplementary Table 4: Type 1 error rates of different tools.
Supplementary Table 5: Comparing LDAK-GBAT and existing tools for the ten UK Biobank phenotypes.
Supplementary Table 6: Reanalysis of the ten UK Biobank phenotypes using only directly-genotyped SNPs.
Supplementary Table 7: Comparing LDAK-GBAT and single-SNP analysis for the ten UK Biobank phenotypes.
Supplementary Table 8: Varying the heritability model for the ten UK Biobank phenotypes.
Supplementary Table 9: Comparing LDAK-GBAT and existing tools for the ICD10, MVP and PGC phenotypes.

Supplementary Table 10: Results of LDAK-GBAT for ICD10, MVP and PGC phenotypes.

### Supplementary Note 1: Mathematical details of LDAK-GBAT

Suppose we have a GWAS of n individuals, and we wish to test a gene containing m SNPs. Suppose the length-n vector $Y$ contains the phenotypic values, the $n \times m$ matrix $X$ contains the SNP genotypes, and the $n \times p$ matrix $Z$ contains covariates (note that for simplicity, we ignored covariates in the main text). For convenience, we assume $Y$ and $X_j$, the jth column of $X$, have been standardized to have mean zero and variance one.

#### The heritability model

When analyzing SNP data, the heritability model describes the prior belief regarding how much each SNP will contribute towards the phenotype. Specifically, it models how $E\left[h_j^2\right]$, the expected heritability of SNP j, varies across the genome. Simple heritability models take the form

$$E\left[h_j^2\right] \propto q_j$$

where the SNP annotations $q_j$ are specified in advance (more complex heritability models replace $q_j$ with multiple sets of SNP annotations [1]. The majority of SNP-based analyses in human statistical genetics use $q_j = 1$; we refer to this as the Uniform Model, because it corresponds to a prior belief that each SNP is expected to contribute equally to heritability [2]. Based on our previous works[1,3-5], we recommend setting $q_j = \left[p_j\left(1 - p_j\right)\right]^{0.75}$, where $p_j$ is the minor allele frequency of SNP j; we refer to this as the Human Default Model.

#### Assumptions of LDAK-GBAT

When testing the association between $X$ and $Y$, LDAK-GBAT assumes

$$Y_i = Z_i\theta + X_i\beta + e_i = Z_{i,1}\theta_1 + \ldots + Z_{i,p}\theta_p + X_{i,1}\beta_1 + \ldots + X_{i,m}\beta_m + e_i$$

where $\theta_k$ and $\beta_j$ are the coefficients for the kth covariate and the jth SNP, respectively, while $e_i$ is the environmental noise for the ith individual. We treat the $\theta_k$ as fixed effects, while the $\beta_j$ and $e_j$ are considered to be random effects, assigned the prior distributions

$$\beta_j \sim N\left(0, q_j\sigma_g^2\right) \quad \text{and} \quad e_i \sim N(0, \sigma_e^2)$$

where $\sigma_g^2$ and $\sigma_e^2$ denote genetic and environmental variances, respectively. Note that because both $Y$ and $X_j$ are standardized, the heritability of SNP j is $\beta_j^2$, which has expected value $q_j\sigma_g^2$. Therefore, the values for $q_j$ determine the heritability model. When $q_j = 1$, LDAK-GBAT assumes the same model as FaST-LMM-Set[6].

**Estimating the variance components**

REML seeks the values of $\sigma_e^2$ and $\sigma_g^2$ that maximize the restricted log likelihood. We start with the full likelihood, whose logarithm is

$$l_F(\sigma_e^2, \sigma_g^2, \theta, \beta) = \frac{-(Y - Z\theta - X\beta)^T(Y - Z\theta - X\beta)}{2\sigma_e^2} - \frac{n}{2}log(2\pi\sigma_e^2) - \frac{\beta^T Q^{-1}\beta}{2\sigma_g^2} - \frac{m}{2}log(2\pi\sigma_g^2) - \frac{1}{2}\sum log(q_j)$$

where $Q$ is a diagonal matrix with entries $q_j$. We integrate the full likelihood across $\theta$ and $\beta$, then log, in order to obtain the restricted log likelihood

$$l_R(\sigma_e^2, \lambda) = \frac{-\gamma}{2\sigma_e^2} - \frac{n-p}{2}log(2\pi\sigma_e^2) + \frac{m}{2}log(\lambda) - \frac{1}{2}\sum log(q_j) - \frac{1}{2}log|Z^TZ| - \frac{1}{2}log|B|$$

where

$$\gamma = Y^TCY - Y^TCXB^{-1}X^TCY, \quad C = I - Z(Z^TZ)^{-1}Z, \quad B = (X^TCX + Q^{-1}\lambda) \quad \text{and} \quad \lambda = \sigma_e^2/\sigma_g^2$$

Note that introduction of $\lambda$ is only for convenience (we find it easier to maximize the restricted log likelihood with respect to $\sigma_e^2$ and $\lambda$, than with respect to $\sigma_e^2$ and $\sigma_g^2$). By differentiating with respect to $\sigma_e^2$, we find that the restricted log likelihood is maximized when $\sigma_e^2 = \gamma/(n-p)$, and therefore our aim is now to find $\lambda'$, the value of $\lambda$ that maximizes

$$l_R\left(\frac{\gamma}{n-p}, \lambda\right) = \frac{-n-p}{2} - \frac{n-p}{2}log\left(2\pi\frac{\gamma}{n-p}\right) + \frac{m}{2}log(\lambda) - \frac{1}{2}\sum log(q_j) - \frac{1}{2}log|Z^TZ| - \frac{1}{2}log|B|$$

We do this using the Newton-Raphson algorithm, where at each iteration, the current value of $\lambda$ is replaced by

$$\lambda - \frac{l'_R}{l''_R} \quad \text{where} \quad l'_R = dl_R\left(\frac{\gamma}{n-p}, \lambda\right)/d\lambda \quad \text{and} \quad l''_R = d^2l_R\left(\frac{\gamma}{n-p}, \lambda\right)/d\lambda^2$$

stopping when the restricted log likelihood changes by less than 0.001 (the user can change this threshold if desired). In order to compute $l'_R$ and $l''_R$, it is necessary to differentiate $\gamma$ and $log|B|$. This is facilitated by first computing the eigen-decomposition $X^TCX = UEU^T$ , as then we have

$$\gamma = Y^TCY - D^T(E + Q^{-1}\lambda)^{-1}D = Y^TCY - \sum D_j^2\left(E_j + \lambda/q_j\right) \quad \text{where} \quad D = U^TX^TCY$$

and

$$log|B| = log|U(E + Q^{-1}\lambda)U^T| = log|(E + Q^{-1}\lambda)U^TU| = log|E + Q^{-1}\lambda| = \sum log\left(E_j + \lambda/q_j\right)$$

both of which expressions can be easily differentiated with respect to $\lambda$.

Note that to calculate a likelihood ratio test (LRT) statistic, we must also maximize the restricted log likelihood under the null model. This is obtained by excluding from the full likelihood all terms corresponding to $X$ or $\beta$, then integrating across $\theta$ and taking the logarithm

$$l_0(\sigma_e^2) = \frac{-Y^T CY}{2\sigma_e^2} - \frac{n-p}{2} log(2\pi\sigma_e^2) - \frac{1}{2} log|Z^T Z|$$

which is maximized when $\sigma_e^2 = Y^T CY/(n-p)$. Therefore, the LRT statistic is

$$S = 2 \times \left[ l_R\left(\frac{\gamma}{n-p}, \lambda'\right) - l_0\left(\frac{Y^T CY}{n-p}\right) \right]$$

**Computing a p-value**

Suppose there are G genes, let $S_g$ denote the LRT statistic for Gene g. Following the approach of FaST-LMM-Set, we assume that $S_g$ has a scaled gamma distribution

$$S_g = f\Gamma(a, b) + (1-f)\delta_0$$

where $f$, $a$ and $b$ are estimated by permutation, as explained below. Thus, the p-value for gene g is

$$P_g = \begin{cases} fP(S_g, a, b), & if S_g > 0 \\ 0.5 + f/2, & if S_g = 0 \end{cases}$$

where $P(S_g, a, b)$ is the probability that a random variable drawn from a $\Gamma(a, b)$ distribution is greater than $S$.

Our permutation procedure for estimating $f$, $a$ and $b$ is almost identical to that used by FaST-LMM-Set. First, we repeat the analysis $K$ times using permuted genotypes (i.e., replacing $X$ with $X[H,]$, where $H$ is a permutation of the integers from 1 to n). Suppose $V$ of the $GK$ permuted LRT statistics are greater than zero. Our estimate of $f$ is $V/GK$ (i.e., the fraction of statistics greater than zero). We then select the $W = min\ (V, GK/10)$ largest permuted LRT statistics (in practice, we have never encountered a scenario where less than 10% of the permuted test statistics are positive, so $W$ always equals $GK/10$). Let $T_1, \ldots, T_w$ denote the selected LRT statistics, ordered so that $T_1$ is the largest. Our estimates of $a$ and $b$ are the values that minimize the mean squared difference between $log\left(P(T_j, a, b)\right)$ and $log\ (E_j)$, where $E_j = (j+1)/(V+1)$ is the expected value of the jth of $V$ draws from a standard uniform distribution.

The small difference between our procedure and that of FaST-LMM-Set is that we generate a new permutation vector $H$ for every gene (i.e., $GK$ times in total), whereas FaST-LMM-Set only generates a new permutation vector at the start of each re-analysis (i.e., $K$ times in total). We chose to update $H$ for each gene because we feel that introducing more randomness might lead to more accurate estimates of the null distribution of $S_g$. However, we note that our change has limited impact on the final p-values (evidenced by Supplementary Figure 1).

**Using summary statistics and a reference panel**

Here we explain how it is possible to compute the LRT statistic for a gene using only summary statistics from single-SNP analysis and a reference panel. The following derivations make four assumptions (which are common to most summary statistic methods):

1 - The single-SNP analysis used least-squares linear regression (note that previous works show that they remain good approximations if the analysis instead used mixed-model linear regression or logistic regression [7,8]).

2 - That $CX_j \approx X_{j}$j and $X_jCX_k \approx X_jX_k$ (note that $CX_j$ is the residual from regressing $X_j$ on $Z$, so these assumptions hold if there is negligible correlation between each SNP and the covariates).

3 - That $1 - Cor\left(CX_j, CY\right)^2 \approx 1$ (i.e., the phenotypic variance explained by any individual SNP is relatively small).

4 – That $(n-p)/n \approx 1$ (i.e., that the sample size is much larger than the number of covariates).

Further, we continue to assume that both phenotypes and SNPs are standardized (this is just for convenience and does not affect the mathematics).

In order to compute the LRT statistic, we must compute the restricted log likelihood and its first and second derivatives. These involve the data ($Y$, $X$ and $Z$) only through the variables $\gamma$ and $B$. Specifically, $\gamma$ and $B$ involve terms of the form $X^TCX$and $X^TCY$ (note that although $\gamma$ also involves the term $Y^TCY$, the term's value is arbitrary, because it is canceled out when computing the LRT statistic).

Our estimate of $X^TCX$ is $nX'^TX'/n'$ , where $X'$ is the matrix of genotypes for the $n'$ samples in the reference panel (standardized so columns have mean zero and variance one). Our estimate of $X^TCY$ is $Z_j\sqrt{n}$. This reflects that when regressing $Y$ on $X_j$ and $Z$, the estimate of the SNP effect size is

$$\hat{\beta}_j = \frac{X_j^TCY}{X_j^TCXj} \approx \frac{X_j^TCY}{n}$$

and the variance of this estimate is

$$Var(\hat{\beta}_j) = \frac{1 - Cor(CX_j, CY)^2}{X_j^{TXj}} \approx \frac{1}{n}$$

It follows that the Wald test statistic from single-SNP analysis is

$$Z_j = \frac{\hat{\beta}_j}{\sqrt{Var(\hat{\beta}_j)}} \approx \frac{X_j^T CY}{\sqrt{n}}$$

Having calculated the LRT statistics for each gene, we must then repeat the analyses using permuted genotypes. This requires us to estimate $X^T CY$, and $X^T CX$, where $X$ now contains permuted genotypes. Our estimate of $X^T CY$ is $nX'^T Y'/n'$ , where $Y'$ is a fake phenotype for the individuals in the reference panel (generated by drawing values from a standard Gaussian distribution). Meanwhile, our estimate of $X^T CX$ remains $nX'^T X'/n'$ (reflecting that permuting the individuals does not affect the correlation between SNPs). Note that because our estimate of $X^T CX$ is unchanged, it is only necessary to eigen-decompose this matrix once (rather than $1 + K$ times), which leads to substantial time savings.

**Implementation details**

Most of the following decisions were made to increase robustness when using summary statistics.

Prior to analyzing each gene, we prune SNPs in high linkage disequilibrium. Specifically, when we encounter a pair, whose correlation squared is above a threshold, we exclude one of the two SNPs at random. By default, the correlation-squared threshold is 0.98 when using individual-level data, or 0.5 when using summary statistics. Supplementary Figure 5 shows that this pruning has limited impact when using individual level data, but can increase power when using summary statistics. We suspect this is because LDAK-GBAT becomes more sensitive to misspecification of $X_j^T X_k$ for pairs of SNPs that are highly correlated.

When analyzing summary statistics, we begin by estimating $R^2$, the proportion of phenotypic variance explained by the gene under least squares regression (i.e., when regressing $CY$ on $CX$). If the estimate of $R^2$ is below 0 or above 1, this suggests errors with the reference panel (e.g., perhaps one or more of the SNPs are poorly genotyped), so we set the LRT statistic for this gene to zero. Similarly, if the value of $\lambda$ that maximizes the restricted log likelihood corresponds to a heritability higher than $10 \times R^2$, we decide the results are unreliable and set the LRT statistic to zero. We recognize that this second check is unsatisfactory (because the choice of 10 is fairly arbitrary). However, we note that across the ten UK Biobank phenotypes, this test never failed when using our recommended reference panel (10,000 UK

Biobank individuals), and in total failed only five times when using 404 non-Finnish, European individuals from the 1000 Genome Project [9].

When maximizing the restricted log likelihood, we start by performing a grid search. For this, we compute the restricted log likelihood for 19 different values for $\lambda$, corresponding to the heritabilities 0, $1\times10^{-6}$, $5\times10^{-6}$, $1\times10^{-5}$, $5\times10^{-5}$, $1\times10^{-4}$, $2\times10^{-4}$, $5\times10^{-4}$, 0.001, 0.002, 0.005, 0.01, 0.02, 0.05, 0.1, 0.2, 0.5, 0.99 and 0.995. We first use this grid search to determine a suitable starting value for $\lambda$ (i.e., we start with the value that resulted in highest restricted log likelihood). We subsequently use it to restrict the search space when performing Newton-Raphson iterations (e.g., if the starting heritability is 0.002, then the Newton-Raphson search is constrained to heritabilities between 0.001 and 0.005, the two adjacent values).

Finally, it is fairly common for summary statistic methods to use shrinkage when using the reference panel to estimate SNP-SNP correlations [10,11]. While LDAK-GBAT offers an analogous feature (e.g., the user can request that the estimate of $X^TCX$ is $0.9 \times nX'^TX'/n'$, instead of $vnX'^TX'/n'$), we find that this has limited effect on the performance of LDAK-GBAT.

**Supplementary Note 2: Existing summary statistic tools for gene-based association testing**

Note that all the following tools require summary statistics and a reference panel, with the exception of sumFREGAT-ACAT, that requires only summary statistics.

MAGMA (multimarker analysis of genomic annotation) computes a gene-based test statistic by summing the chi-squared test statistics from single-SNP analysis. A p-value is obtained by comparing the gene-based test statistic to its expected distribution under the mean (in the most recent version of MAGMA, the null distribution is assumed to be mixture of chi-squared distributions) [12].

GCTA-fastBAT (genome-wide complex trait analysis fast set-based association test) computes a gene-based test statistic by summing the chi-squared test statistics from single-SNP analysis (the same as MAGMA). A p-value is obtained via permutations [13].

sumFREGAT (Fast Region-Based Association Tests on Summary Statistics) implements a variety of gene-based association test tools, of which we use ACAT, SKAT-O, and PCA [14,15].

ACAT computes a gene-based test statistic by first converting p-values from single-SNP analysis to Cauchy variables, then taking a weighted sum. A p-value is obtained analytically.

STAT-O (optimized SNP-set (sequence) kernel association test) uses a random-effects linear model (the same as LDAK-GBAT), where the prior distributions for effect sizes depend on the minor allele frequency (MAF) of SNPs. A p-value is obtained via a score test.

PCA (principal component analysis) use a fixed-effect linear model, where the predictors are the top principal components (i.e., linear combinations of genotypes).

Note that we briefly considered MLR and FLM, two other tools within sumFREGAT. However, our analysis of permuted phenotypes (Supplementary Table 4) indicated that these tools have inflated type 1 error rates (e.g., the number of genes with $P \leq 1 \times 10^{-6}$ was approximately 1000 times higher than expected if the tools were well-calibrated), and hence we excluded these from subsequent analyses.

**Supplementary Note 3: Further details of data**

**Ten UK Biobank phenotypes**

The ten phenotypes are body mass index (data field 21001), college education (6138), forced vital capacity (3062), height (50), hypertension (20002), impedance (23106), neuroticism score (20127), preference for evenings (1180), pulse rate (102) and systolic blood pressure (4080). Note that these are a subset of the 14 we used in our previous work [2] (we excluded difficulty falling asleep, ever smoked, reaction time and snorer because for these there were no significant genes from either gene-based association testing or single-SNP analysis). For each phenotype, we had between 220,398 and 253,313 unrelated, white British individuals. When performing gene-based association testing, we used only 50,000 individuals (picked randomly for each phenotype); however, for single-SNP analysis, we used up to 200,000. We computed summary statistics for each of the 7,186,768 SNPs in our primary reference panel (see below) by performing classical linear regression (including for the two binary phenotypes, college education and hypertension), including 13 covariates (age, sex, Townsend Deprivation Index and ten principal components).

**72 ICD10 Phenotypes**

In total, the UK Biobank details 19155 ICD10 codes (data field 41202) that span four levels. We restricted to the 349 Level 2 codes with prevalence at least 1% (note that for some codes, over 80% of cases were male or female; for these, we excluded the less-common sex when computing prevalence and for all subsequent analyses). For each of the 349 codes, we constructed a GWAS using all $n_A$ cases and $n_U$ randomly-picked controls, where $n_U = min\ (10n_A, 50000)$. We used REML to estimate SNP heritability (assuming the LDAK-Thin Model) [1,3]. Then we selected the 72 phenotypes with most significant SNP heritability (Z-statistic >10), and computed summary statistics as described above (i.e., using classical linear regression including 13 covariates). Supplementary Table 1 provides details for each phenotype.

**18 Million Veterans Project and nine Psychiatric Genomics Consortium phenotypes.**

Summary statistics for Million Veterans Project and Psychiatric Genomics Consortium phenotypes were acquired from the respective consortia and are detailed in Supplementary Tables 2 and 3. If a GWAS provided information scores, we excluded SNPs with score below 0.8.

### Reference panels

Our primary reference panel comprises 10,000 unrelated UK Biobank individuals genotyped for 7,186,768 SNPs with MAF ≥ 0.01 and imputation information score ≥0.8. We ensured the 10,000 individuals were distinct from the 200,000 individuals used for each of the ten UK Biobank phenotypes (however, there will be by-chance overlap with the individuals used for the ICD10 GWAS).

For Supplementary Figure 3, we constructed a reference panel using 404 non-Finnish, European individuals, recorded for the same 7,186,768 used in our primary reference panel.

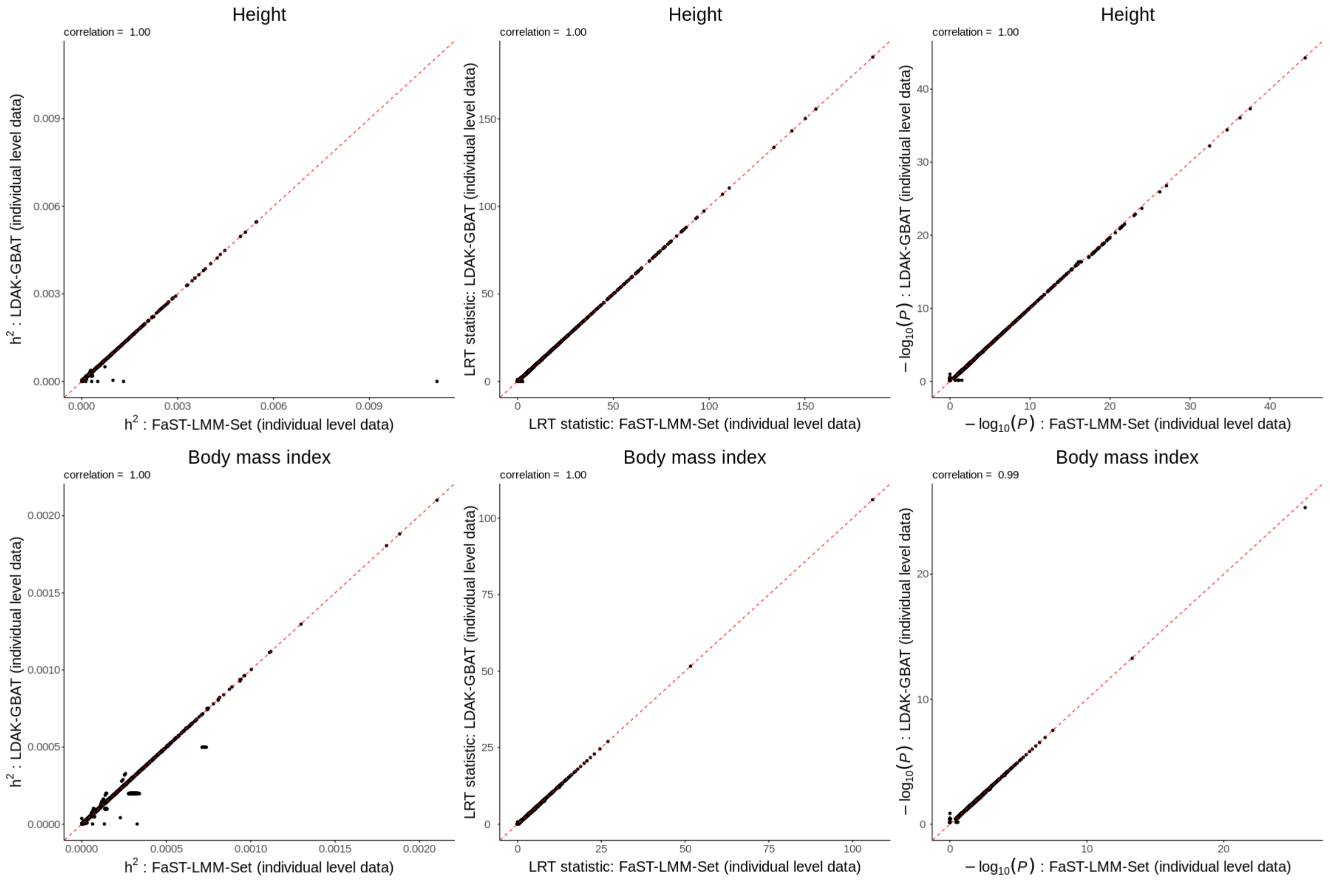


**Supplementary Figure 1: Comparison of FaST-LMM-Set and LDAK-GBAT.**
For the UK Biobank phenotypes height (top row) and body mass index (bottom row), we compare the estimated heritabilities (left), likelihood ratio test statistics (middle) and -$\log_{10}(P)$ (right) from FaST-LMM-Set (x-axis) and LDAK-GBAT (y-axis). To enable a fair comparison, we run LDAK-GBAT using individual-level data, assuming the Uniform Model and do not prune genes (i.e., so its model matches exactly that used by FaST-LMM-Set). In general, we see almost perfect concordance (and although not shown, this is also the case for the remaining eight UK Biobank phenotypes). Although there are small differences between estimates of heritabilities, they occur only for non-significant genes, and we expect these reflect minor details in the implementations of each tool (for example, while both tools estimate heritability using Newton Raphson iterations, LDAK-GBAT decides the starting values based on a grid search, whereas FaST-LMM-Set sets it agnostically).

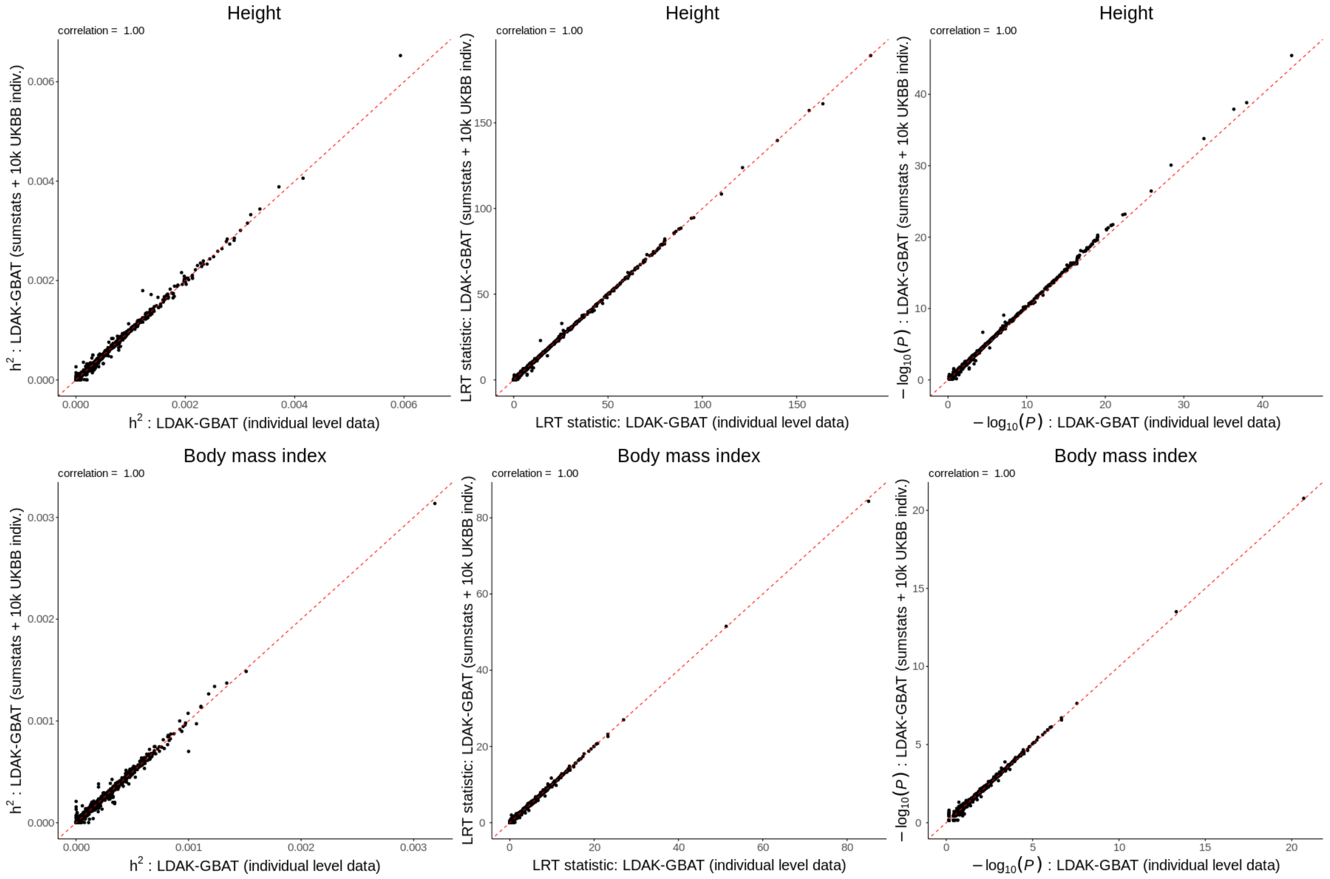


**Supplementary Figure 2: Comparison of LDAK-GBAT using individual-level data and summary statistics**.

For the UK Biobank phenotypes height (top row) and body mass index (bottom row), we compare the estimated heritabilities (left), likelihood ratio test statistics (middle) and -$\log_{10}(P)$ (right) from LDAK-GBAT using individual level data (x-axis) or using summary statistics (y-axis). We run LDAK-GBAT assuming the Human Default Model and using the default pruning of genes (when analyzing individual-level data, LDAK-GBAT ensures no pair of SNPs in a gene has squared correlation above 0.98; when analyzing summary statistics, the threshold is reduced to 0.5). In general, we see high concordance (and although not shown, this is also the case for the remaining eight UK Biobank phenotypes), indicating that it is feasible to test genes for association using only summary statistics.

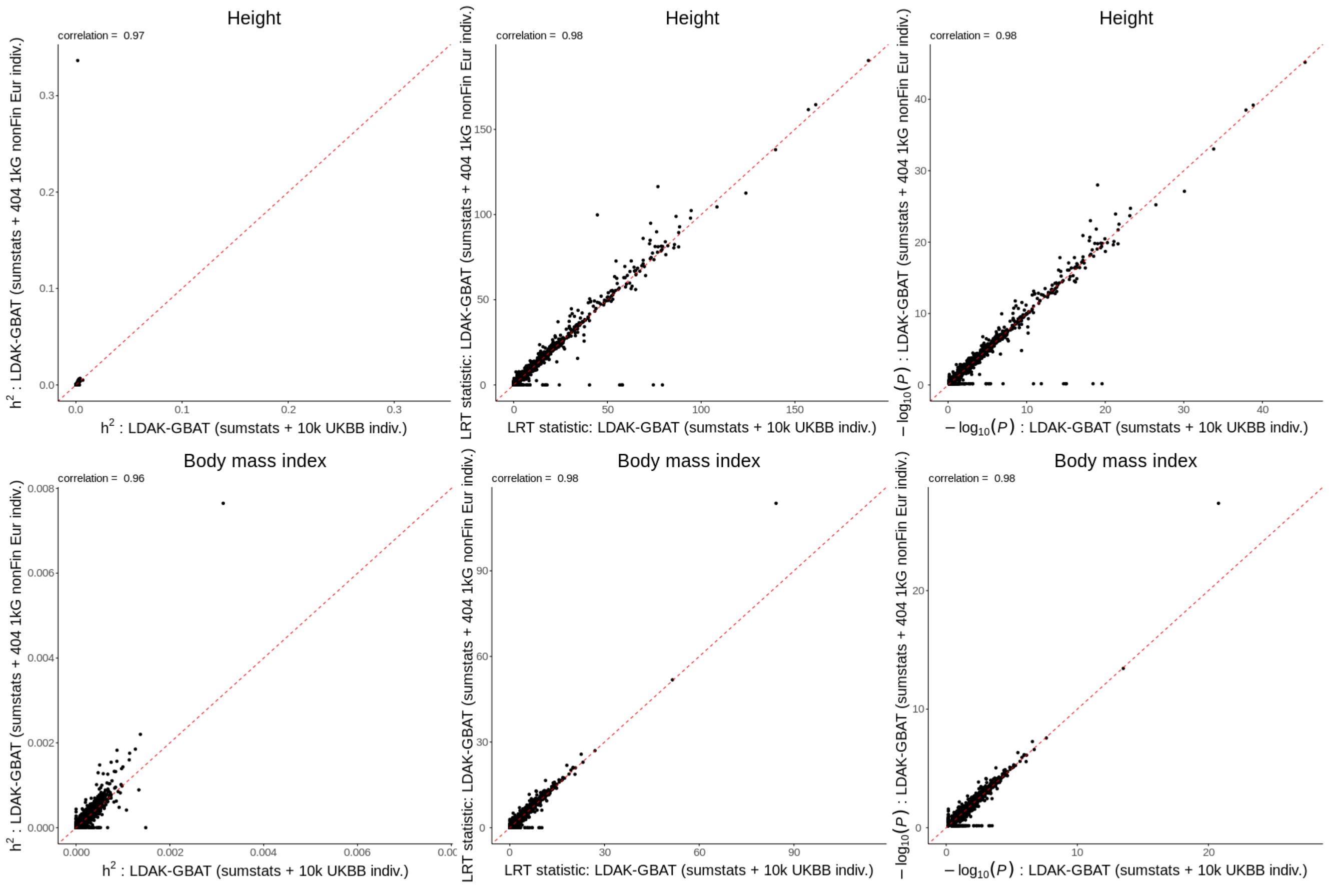


**Supplementary Figure 3: Impact of changing the reference panel.**
For the UK Biobank phenotypes height (top row) and body mass index (bottom row), we compare the estimated heritabilities (left), likelihood ratio test statistics (middle) and -$\log_{10}(P)$ (right) from LDAK-GBAT using as a reference panel 10,000 individuals from the UK Biobank (x-axis) or 404 non-Finnish, European individuals from the 1000 Genomes Project reference panel (y-axis). In general, we see good concordance (and although not shown, this is also the case for the remaining eight UK Biobank phenotypes). However, we note that power is slightly reduced when using the 1000 Genomes Project reference panel (for example, there are eight genes that are no longer significant for height when switching from the UK Biobank reference panel). This reflects that LDAK-GBAT benefits from use of a large reference panel (e.g., at least 2000 individuals) that is very closely matched (with respect to ancestry) to the GWAS from which the summary statistics were obtained.

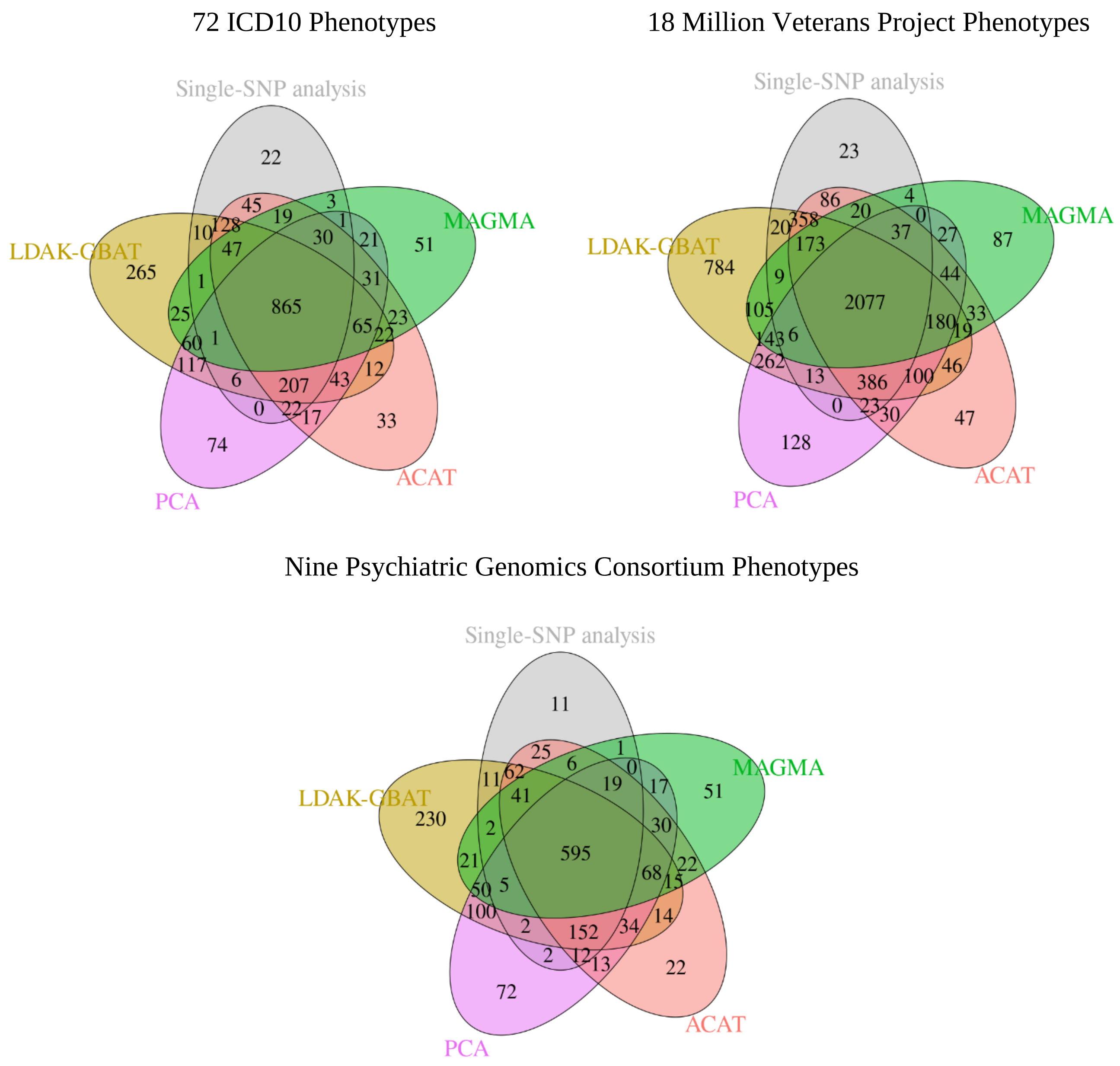


**Supplementary Figure 4: Overlap between significant genes from different tools.**
The Venn Diagrams show the overlap between the significant genes found by LDAK-GBAT, MAGMA, sumFREGAT-PCA, sumFREGAT-ACAT and single-SNP analysis, for the three groups of phenotypes. For example, the first Venn Diagram shows that across the 72 ICD10 phenotypes, LDAK-GBAT found 1874 significant genes, of which 865 were also found by all four other tools, 60+117+1+6=184 were also found by sumFREGAT-PCA, while 265 were not found by any other tool.

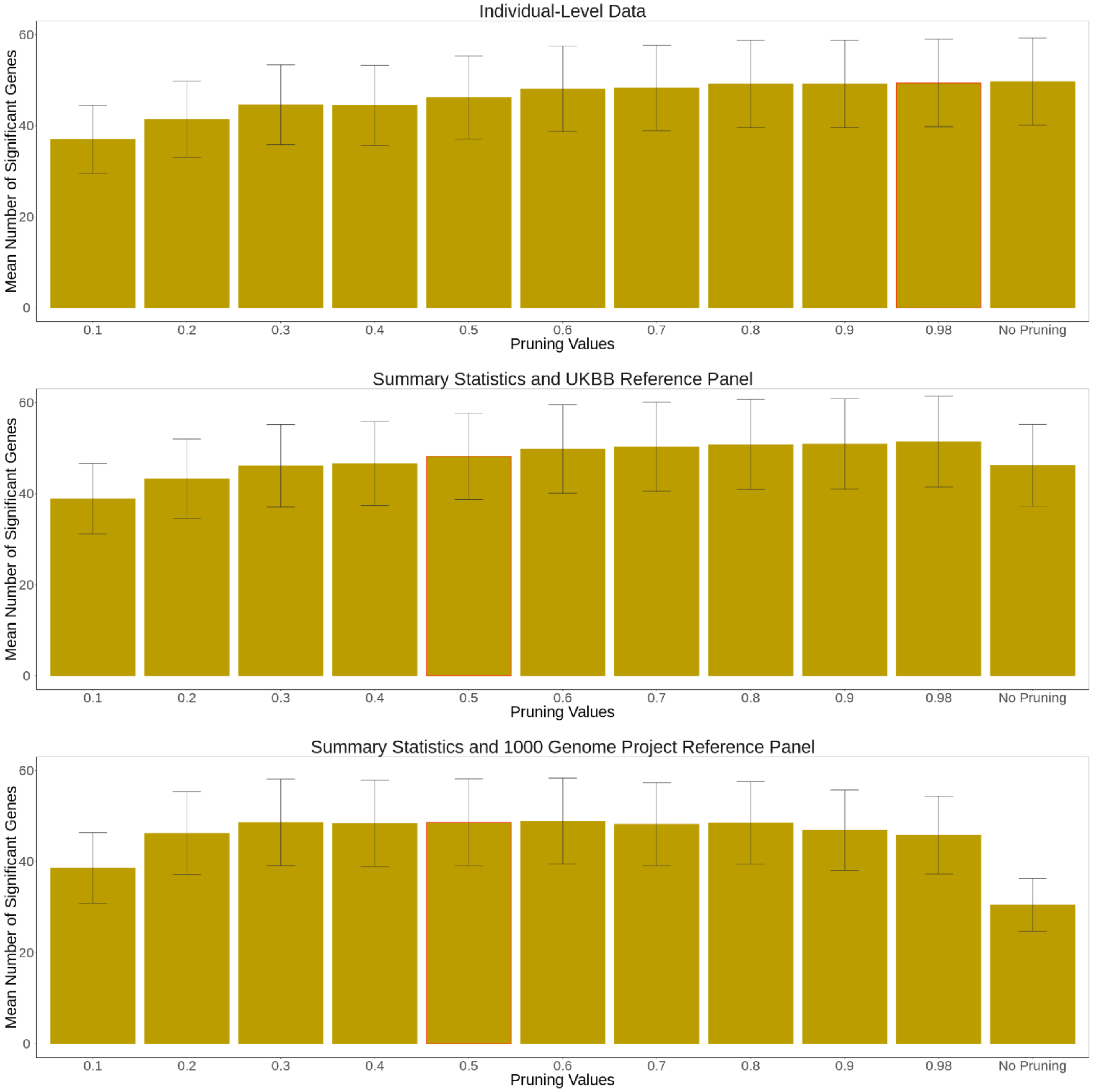


**Supplementary Figure 5: The impact of pruning genes.**

Prior to analyzing each gene, LDAK-GBAT prunes its SNPs so that no pair remains with squared correlation above a threshold. We compare the mean number of significant genes ($P \leq 2.8 \times 10^{-6}$) across the ten UK Biobank phenotypes for different pruning thresholds. We see that when using individual level data (top plot), pruning has limited impact. When using summary statistics with a reference panel of 10,000 UK Biobank individuals (middle plot), it is beneficial to have light pruning. When using summary statistics with a reference panel of 404 non-Finnish, European individuals from the 1000 Genome Project (bottom plot), it is beneficial to have moderate pruning. Based on these results, we set the default threshold to 0.98 when analyzing individual-level data (although pruning does not appear to increase power, removing very highly correlated SNPs will reduce the computational demands), and to 0.5 when analyzing summary statistics (this potentially makes LDAK-GBAT more robust when using smaller and/or less well-matched reference panels).

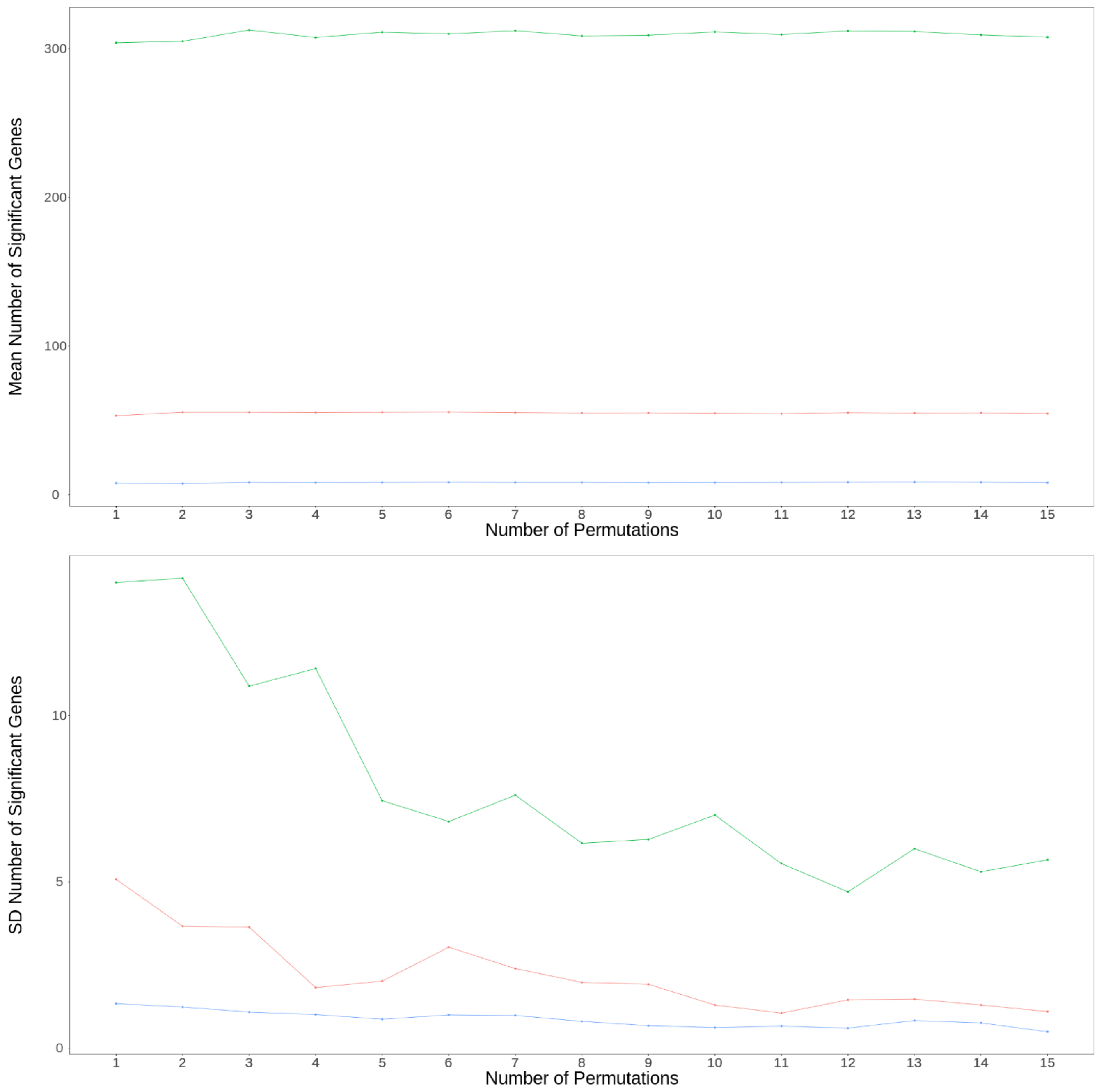


**Supplementary Figure 6: Impact of number of permutations.**
LDAK-GBAT uses permutations to obtain gene-based p-values. We analyze the UK Biobank phenotypes forced vital capacity (red), height (green) and systolic blood pressure (blue) twenty times each, and compare how the mean (top plot) and standard deviation (bottom plot) of the number of significant genes varies with the number of permutations. Based on these results, we set the default number of permutations to ten. This is because although increasing the number of permutations produces more robust results (i.e., the number of significant genes varies less across replicates), there appears limited benefit to using more than ten permutations (because the standard deviation reaches a plateau).
